# Computational Causal Discovery for Posttraumatic Stress and Negative Self Image in Young Maltreated Children

**DOI:** 10.1101/2022.07.20.22277681

**Authors:** Glenn N. Saxe, Leah J. Morales, Sisi Ma, Mehmet Urgurbil, Constantin Aliferis

## Abstract

**Objectives:** This article features the application of computational causal discovery (CCD) methods to determine the mechanism for Posttraumatic Stress (PTS) in young, maltreated children, in order to advance knowledge for prevention. Advances in prevention require research that identifies causal factors, but the scientific literature that would inform the identification of causes are almost exclusively based on the application of correlational methods to observational data. Causal inferences from such research will frequently be in error. We conducted the present study to explore the application of CCD methods as an alternative – or a supplement – to experimental methods, which can rarely be applied in human research on causal factors for PTS.

**Methods:** A data processing pipeline that integrates state-of-the-art CCD algorithms was applied to an existing observational, longitudinal data set collected by the Consortium for Longitudinal Studies in Child Abuse and Neglect (LONGSCAN). This data set contains a sample of 1,354 children who were identified in infancy to early childhood as being maltreated or at risk.

**Results:** A causal network model of 251 variables (nodes) and 818 bivariate relations (edges) was discovered, revealing four direct causes and two direct effects for PTS at age 8, within a network containing a broad diversity of causal pathways. Specific causal factors included stress, social, family and development problems: and several of these factors point to promising approaches for prevention.

**Conclusions:** These results indicate that CCD methods show promise for research on the complex etiology of PTS in young, maltreated children.

## Introduction

This article features the application of computational causal discovery (CCD) methods to determine causal factors for Posttraumatic Stress (PTS) in young, maltreated children. 3.6 million reports of child maltreatment were submitted to child protection authorities in the United States in 2014 ^1^ and approximately 30-40% of maltreated children will acquire PTS.^2-4^ A recent metaanalysis on the literature reported that 25% of children exposed to interpersonal trauma, acquire PTSD.^5^ PTS has been associated with significant impact on neurodevelopment, adaptive functioning, and physical and mental health. ^6-13^ Accordingly, there is a great need to prevent PTS in maltreated children. Unfortunately, few preventative interventions are available for PTS in maltreated children, and the results of preventative interventions for PTS for other types of trauma, have been modest.^14-19^

Advances in prevention requires research that identifies causal factors,^20,21^ but the scientific literature that would inform the identification of causes are almost exclusively based on the application of correlational methods to observational data. In the pages of this journal, we have recently described the endeavor to infer causes from observational correlational data as *navigating in a causal labarynth*.^22^ Causal inferences from such research will frequently be in error.^20,21^ Experimental research can infer causes, but such research – for all practical purposes – cannot be conducted with humans (e.g. randomizing a child to a trauma condition). Thus, advances in prevention will need to come from the establishment of alternative methods to experimental research to identify causal factors for PTS. We conducted the present study to explore the application of CCD methods for such a purpose.

It is beyond the scope of this article to review in detail the literature on causal inference. These matters have been extensively investigated elsewhere, including both rigorous mathematical proofs and extensive empirical validation studies using methods to discover causes using gold standard causal discovery data sets where true causes are known.^20-27^ These methods have also yielded important advances in non-psychiatric medical fields and have been successfully applied in psychiatric research, to a large extent by our group.^28-36^ Although there are strong reasons to believe that application of CCD methods to psychiatric research can lead to similar advances, it must be acknowledged that these methods have hardly been applied to psychiatric data.

Accordingly, findings from application of CCD methods in psychiatry should be appraised with a commensurate level of scrutiny. The present study is, to the best of our knowledge, the first to apply CCD methods to discover causal factors for PTS in maltreated children.

Since most readers are likely unfamiliar with CCD methods and how they enable causal inference, we provide a brief non-technical description, next: with more detail provided in Supplement 1.

### Causal Inference from Observational Data

We start by providing a definition that explicitly or implicitly is used by all state-of-the-art CCD algorithms and is consistent with what is considered as causal in biomedicine:^20,21^

#### A. Defining direct and indirect causation

##### Definition 1 (Operational criterion for causation)

Assume that a variable A can be manipulated by a hypothetical experimenter to take values, each one denoted as a_i_. Assume also that the experimenter can manipulate A (e.g. give a drug to a patient or not, where A stands for type of treatment received). We denote the manipulation of A by the experimenter to take value aj, as: do(A=aj). If the experimenter assigns values to A according to a uniformly random distribution over values of A, and then observes P(B|do(A = a_i_))≠P(B|do(A = a_j_)) for some i and j, (and within a time window *dt*), then variable A is a cause of variable B (within *dt*). Intuitively, P(B|do(A = a_i_))≠P(B|do(A = a_j_) means that manipulating A (i.e. do(A = a_i_) or do(A = a_j_)) results in a different distribution of variable B in the manipulated population.

##### Definition 2 (Direct and indirect causation)

Assume that a variable A is a cause of variable B according to the operational criterion for causation in definition 1. A is an indirect cause for B with respect to a set of variables V, if and only if A is not a cause of B for some assignment (by manipulation) of values of V - {A, B}, otherwise A is a direct cause of B. Variables representing direct causes serve to *mediate* the relationship between an indirect cause and the effect.

#### B. Mapping of data to the causal process generating the data (i.e. inferring causes from observational data)

Notice that the above definition hinges on experimental data. To infer causality from observation data, we need to map observation data to the causal process that generates the data. The existence of a set of causal relationships of the type “A causes B” in the vast majority of distributions (so-called “Faithful” distributions) guarantee that specific conditional dependencies and independencies will be observed in the data in a way that directly corresponds to the causal mechanisms of the process that generates the data. For example, consider the situation that A directly causes B, B directly causes C, and no other direct causal relationship exists among them, and that the distribution is Faithful. In this case, all variables are mutually associated. Also, A will become independent of C, if we condition on B (i.e., knowing the values of B blocks the association/dependency/information transfer between A and C). This is an application of the **Causal Markov Condition (CMC)**, a distributional assumption that allows us to infer all statistical independecies observed in data, if we know the causal process generating the data. The distributional assumption of the **Causal Faithfulness Condition (CFC)** says that *all* indepedencies in the data are represented by the causal graph representing the causal process, combined with the CMC. If the CMC and the CFC hold, it follows that all dependencies and independencies in the data correspond precisely to (i.e., form a perfect map of) the true causal structure that generated the data. In other words, an algorithm (or analyst if the number of variable is really small) can measure dependencies and independencies in the data and correctly infer the existence or non-existence of precise direct and indirect causality, without conducting experiments. The combination of CMC+CFC constitutes the aforementioned distribution assumption of Faithfulness. How common are faithful distributions? The answer is that the non-faithful distrbutions among all possible distributions are exceedingly rare (Lebesque measure 0)^37^ and, of the few cases where faithfulness is violated, special algorithms are available that can help manage this problem.^38^

#### C. Discovering the Most Parsimoneous Causal Relations in the Data

A particularly powerful discovery tool – and one centrally related to the methods we apply – concerns the Markov Boundary of a target outcome variable. The Markov Boundary provides the smallest set of variables that achieves maximum possible predictive accuracy of the target variable (most parsimonious prediction) under broad assumptions about classifiers used, data distribution, and error metrics of interest. The Markov Boundary not only enables *most parsimonious prediction*, it also possesses a *causal function* since, in the majority of distributions, and when confounding is restricted, it includes the direct causes, direct effects and direct causes of the direct effects of the target variable. ^20,21,23,24^

### Causality and Disorders of Complex Etiology

Psychiatric disorders such as PTSD almost certainly have complex etiologies.^8,35,39,40^ For such disorders, it is expected that there will be a very large number of interacting causal pathways (of linked indirect and direct causes) for expression of the disorder. Such a state-of-affairs is a hallmark of complex systems (and may explain why complex disorders can be so hard to prevent or to treat). However, knowledge about the nature of complex systems may also reveal unique opportunities for intervention. In complex systems, a small number of causes will influence a large proportion of causal pathways, and most causes influence a small proportion of pathways.^41,42^ We integrate methods designed to leverage this opportunity by detecting those causal variables with influence over the greatest proportion of pathways into the target variable. We emphasize that contrary to the methods designed to discover specific direct and indirect causes to a target outcome variable – that provide guarantees for correctness under well specified conditions – network-connectivity approaches for determining causal variables with broadest influence, are presently a heuristic strategy: This heuristic, however, has the advantage that it describes higher-level properties than methods designed to only infer direct and indirect causes. In the research described next, we apply a specific protocol – called the *Protocol for Computational Causal Discovery in Psychiatry (PCCDP)* – to implement the causal discovery methods just described, to observational data sets used in the field. This protocol was applied to discover a complex network revealing the causes and effects of PTS in maltreated 8 year olds. Of note: PCCDP is an updated version of a previously published protocol called the Complex Systems-Causal Network (CS-CN) method.^35^

## Methods

### Participants

This study analyzed a data set from the Consortium for Longitudinal Studies in Child Abuse and Neglect (LONGSCAN), comprising a sample of 1,354 children identified as being maltreated or at risk. Children and families underwent comprehensive assessments from age 4, with reassessment every two years. Because this study focused on the onset of PTS in early childhood, and with PTS assessment introduced at 8 year-old wave, we included information from the 4, 6, and 8 year-old waves. For a full description of recruitment and data collection procedures see.^43^ The study protocol was approved by NYU Langone Health IRB.

### Procedure

PCCDP was applied to the LONGSCAN data set, and this protocol follows a specific set of steps integrating prior knowledge in the process of CCD, to discover specific causes and effects of psychiatric disorders of interest. These steps are described next, and illustrated in Figure 1.

**Figure 1:**
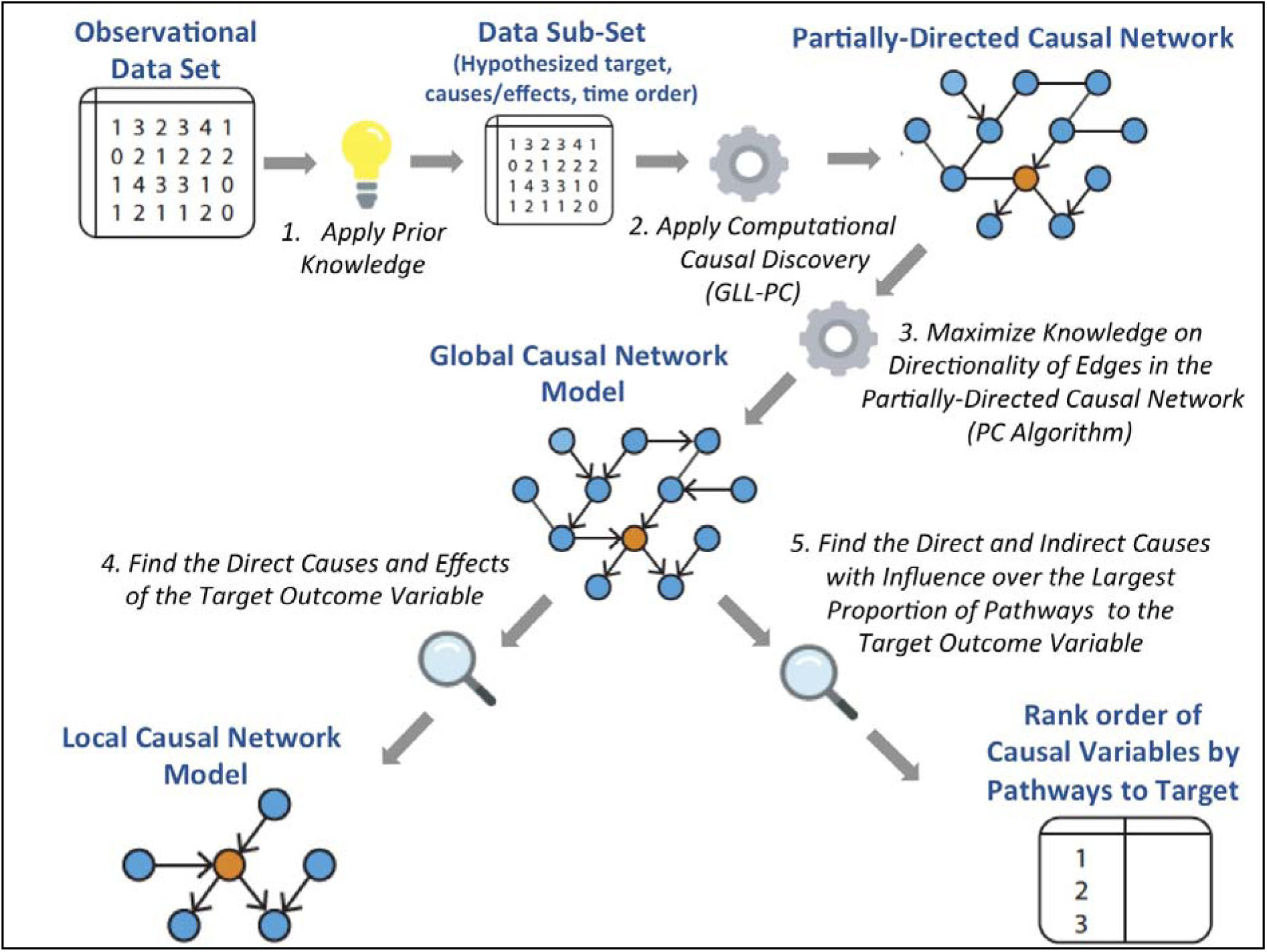
The Protocol for Computational Causal Discovery in Psychiatry (PCCDP)

#### 1. Apply Prior Knowledge to an Observational Data Set to Create a <u>Data Subset</u> based on Hypothesized Causes and Effects of a Designated Target Outcome Variable

As illustrated in Figure 1, PCCDP allows investigators to apply prior knowledge about the phenomenon under study to identify variables and relations considered for analysis. Broadly, PCCDP considers two types of prior knowledge to establish a Data Subset of an observational data set, for computational analysis:

##### i. Prior knowledge on the **variables to include** in the analysis

Although, an important advantage of the algorithms used within PCCDP is their capacity to consider large numbers of variables in samples of modest size, thus minimizing the considerable risk of missing direct and indirect causes as well as failing to analyse hidden common causes that would confound the results of causal discovery. It is also advantageous to be guided by the literature in one’s field, to exclude variables from consideration: a. When there is no reason to believe the variable would have any relation to the target (e.g. For PTS target: child’s color preference, brand of family car). b. When multiple variables measure the same construct (e.g. a data set contains four similar variables measuring depression). This first application of prior knowledge (i.e. variable consideration) seeks to constrain the introduction of superfluous information into the process of causal modeling, and minimize potential violations of faithfulness.

##### ii. Prior knowledge on the **relationship between variables**, such as

i. time order between variables based on knowledge of time of measurement or presumed time of effect or, ii. causal relationships that have been established from prior experimentation. This second application of prior knowledge seeks to leverage *known facts* about relationships, in the process of causal modeling (e.g. fact that causal relations cannot go backwards in time; fact that experiments can determine causal relations).

In the present study, prior knowledge was applied by searching LONGSCAN data set to select variables/relations as follows:

###### a. Selection of the outcome target

LONGSCAN data set was reviewed for variables that best measured PTS. Posttraumatic Stress T-score was chosen as the target outcome variable given its demonstrated strong psychometric properties.^44^ PTS T-score was derived from Trauma Symptom Checklist ^44^ administered during the 8 year wave (Posttraumatic Stress T-Score: M = 52.92, SD = 12.67), (age in months: M= 99.1, SD = 4.9). Of the 1,354 subjects in the data set, 1,053 had data related to this variable and, accordingly, data from these 1,053 subjects were included in the analysis.

###### b. Selection of hypothesized cause and effect variables

Two-hundred and fifty-two variables were pre-selected based on their plausibility as causes or effects of PTS, and information equivalency with included variables. Such variable pre-selection was guided by prominent theory in the field.^8,45^ Variables were chosen to cover a diversity of modalities including demographics, perinatal and developmental factors, socio-environmental context, cognitive and behavioral factors, and Child Protective Services (CPS) records.

###### c. Designation of time epoques for measured variables

The data set contained information from the subject’s prenatal period to data collection at 8 years old in 3 waves at 4, 6, and 8 years old. Based on how the data was collected, and the data set organized, eight time epoques were defined: 1. Constitutional (e.g. age, sex), 2. Perinatal, 3. 0-4 years, 4. 4 years, 5. 4-6 years, 6. 6 years, 7. 6-8 years, 8. 8 years.

The resulting Data Subset was then input for computational processing, along with a defined Variable Table, describing each variable, its numeric type (e.g. ordinal, cardinal) and its time époque. *Importantly, no discretion over the processing of variables was allowed, once the Data Subset and Variable Table is entered as input*.

Each variable in the Data Subset can be found as is in the original LONGSCAN data, except for ten variables which were computed from other variables for purposes of the present study.^43^ Variables were also renamed for sake of clarity and graph visualization. The list of variables and corresponding original names (or names of original variables from which they were constructed) and designated time epoques are provided in Supplement 2.

#### 2. Apply Computational Causal Discovery Methods to the Data Subset to Determine a Partially-Directed Causal Network

Causal edge detection was accomplished by integrating a well-validated set of algorithms called GLL-PC ^23,24^ which discover causal edges by using tests of conditional independence among variables in the Data Subset with the semantics/definition of a causal edge offered in the Introduction.^20,21^ GLL-PC employs local causal discovery and is followed by local-to-global algorithms from best-of-class Local to Global Learning (LGL) families.^20,21,23,24,25,26^ As shown in Figure 1, GLL-PC processes the Data Subset to create a causal network via inference of all causal relationships consistent with the data. In addition to identifying causal relationships, the inferred network can also be used for optimal predictive modeling via elicitation of the Markov Boundary for each of the variables. ^23,24,25^ Because GLL-PC was provided with time epoque information, it was also able to provide direction between many of the variables. Within PCCDP, GLL-PC was programmed to draw direction from variables forward in time (when time order is known). This procedure yields a <u>Partially Directed Causal Network</u> of nodes (variables) and the edges (bivariate causal relations) that connect them. GLL-PC was applied with the following fixed parameters: a *max-k* (maximum conditioning set size) of 3, and a *p-val*ue threshold of 0.05.

#### 3. Maximize Knowledge on Directionality of Causal Edges to Determine the <u>Global Causal Network Model</u> of PTS

To orient any remaining undirected edges (i.e. edges between variables from same time epoque), PCCDP applies techniques derived from the PC algorithm.^21^ Using “sepsets” from the conditional independence tests performed by GLL-PC, PC’s edge orientation rules are employed to orient Y-structures (e.g., A → C ← B) and propagate the orientation of appropriate edges in the given undirected or partially-directed causal network. Observational data can only resolve causal directions up to the Markov Equivalence class, i.e. the set of causal graphs that share same edge orientations consistent with the data, leaving the rest as unknown.^21^ As illustrated in Figure 1, this procedure results in the <u>Global Causal Network Model</u> of PTS.

#### 4. Find the Direct Causes and Effects of the Target Outcome Variable to Determine the <u>Local Causal Network Model</u> of PTS

The previous computational steps create a Global Causal Network Model based on observed causal influences between all variables in the Data Subset. This Global Causal Network Model contains the variables within what is called the <u>local causal neighborhood</u> of the target outcome variable (e.g. PTS), and also the local causal neighborhoods of all variables in the Data Subset (in order to learn pathways of sequences between indirect and direct causes to the target outcome variable). PCCDP also reveals the Markov Boundary for each variable since the latter comprises the direct causes, direct effects, and direct causes of the direct effects of each variable, including PTS. This <u>Local Causal Network Model</u> for PTS is one the two main ‘outputs’ of PCCDP, shown in Figure 1.

#### 5. Find the <u>Direct and Indirect Causes with Influence Over the Largest Proportion of Pathways</u> to PTS

In general, once the global causal graph is discovered, a variety of graph search algorithms combined with quantitative causal inference can be used to derive sets of variables that separate a cause of PTS from PTS or that achieve a desired degree of influence on PTS. Such calculations are very expensive and even prohibitive for large, complex networks, however. In the present study, we instantiate PCCDP with a heuristic strategy as follows: In order to determine the causal variables that have influence over the largest proportion of pathways to the target, a subnetwork of all pathways in the Global Causal Network was examined that terminate at the designated target variable, restricted to four path-lengths or less (to exclude variables with distal influence on target). PCCDP evaluates (heuristically) the effect of intervention on causal variables by determining the number of intact pathways to the target that become eliminated, when each causal variable is deleted. As shown in Figure 1, PCCDP produces, as output, a rank order of causal variables by proportion of pathways to the target, eliminated, when specific causal variables are deleted.

##### Additional Safeguards

###### 1. Supporting Faithfulness Assumption through Multiplicity Testing

PCCDP includes a safeguard called <u>Multiplicity Testing</u> to ensure that the data contains a single Markov Boundary (and local casual neighborhood) for the outcome of interest.^38^ Multiplicity refers to the presence of multiple Markov Boundaries for a given target variable and is a form of faithfulness violation. If Multiplicity is present, despite efforts to include variable sets that are not equivalent, causal discovery algorithms that assumes single Markov Boundary return incomplete and erroneous information (may miss important causal variables, and mistake irrelevant variables as causally important). Multiplicity Testing was implemented with the TIE* algorithm.^38^

###### 2. Determining Stability of Causal Edges through Bootstrapping Procedures

PCCDP integrates a bootstrapping analysis to determine the stability of the resulting Global Causal Network Model with respect to sampling variability. The Data Subset was bootstrapped 100 times and Global Causal Network Models were generated for each iteration. The stability of an individual edge was calculated as the percent (%) of bootstrapped networks it was detected.

### Software and tools

PCCDP was programmed in MATLAB R2016a.^46^ Post-analysis network visualization was carried out in Cytoscape 3.6.1.^47^ Network pathway analysis was conducted in part using Cytoscape plugin, Pathlinker.^48^

## Results

A Global Causal Network Model of 251 nodes and 818 edges was detected (1 node was isolated from the main component of the graph). Of the total edges, 290 (35.5%) were oriented based on time époque and 485 (59.3%) by the PC edge-orientation rules. In total, 775 (94.7%) edges were oriented and 43 (6.3%) edges remained unoriented.

The complete edge list and edge stability estimated from bootstrapping analysis, is provided in Supplement 3.

### Direct Causes and Effects of PTS (Local Causal Network Model)

In Figure 2, a visualization of variables found within the Markov Boundary for PTS at age 8 (i.e. the Local Causal Network Model) is presented. These variables reflect the minimal set of information required to predict the value of the target available in the dataset. Conditioned on these nodes, PTS T-score is rendered independent of the rest of the nodes in the network. The visualization shows the direct causes, direct effects, and direct causes of the direct effects of the target (i.e. target’s Markov Boundary). This network model contains 19 nodes and 28 edges, and is configured in hierarchical layout to best visualize the flow of causal influence. As can be seen, there are four direct causes to PTS: Emotional Maltreatment Records (age 0-4), Physically Assaulted (age 8), Feels Safe (age 8), and Witness Violence (age 8). There are two paths from PTS (i.e, direct effects): Severe Assault: Non-Caregiver (age 8), and Negative Self Image (at age 8). There is one undirected edge with Physically Threatened (age 8) because PCCDP was unable to determine tdirection of this relationship.

**Figure 2.**
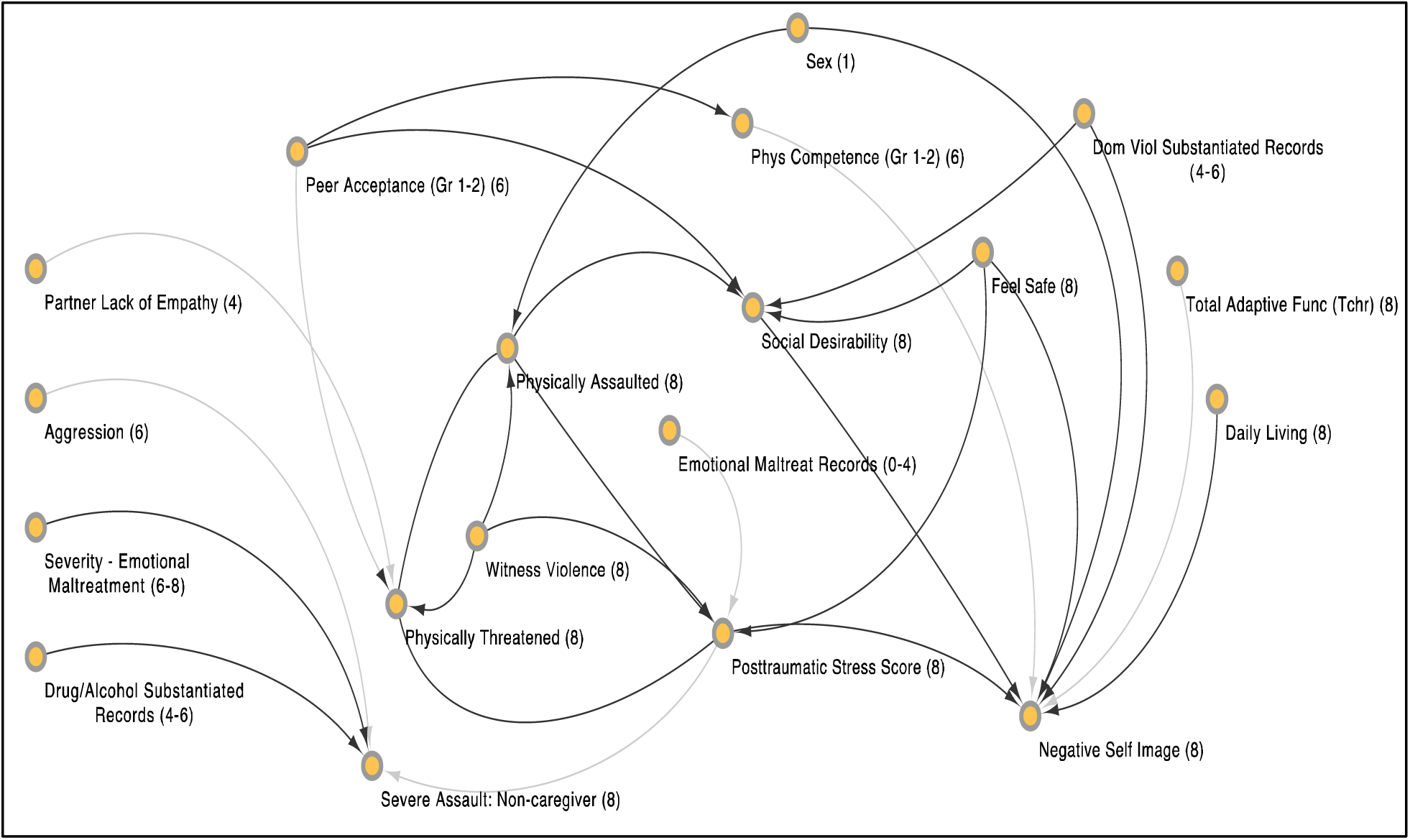
Network visualization of the Markov Boundary of Posttraumatic Stress T-score at age 8. The numeral in parenthesis within each node indicates the time époque (1-8) of the variable represented by the node (which indicate the child’s age when the variable was measured and/or exerted its effect). Some edges are shaded in a lighter gray. This designates edges that appeared in less than 30% of bootstrapped samples. These edges are considered less stable than the other edges in the proximal causal network. This network visualization was generated using Cytoscape 3.6.1.

The test for multiplicity revealed that the observed Markov Boundary is unique for PTS, strengthening the causal validity of the Local Causal Network Model by supporting the assumption of local faithfulness.

### Direct and Indirect Causes influencing the Largest Proportion of Paths to PTS

We examined all paths (of four path-lengths or less) in the Global Causal Network that terminated at PTS target variable. One hundred and thirty-eight unique paths were found. Many nodes are part of multiple paths. We heuristically evaluated the effect of intervention on each node by computing the proportion of paths eliminated when a node was deleted from the network. Figure 3 shows the 10 nodes that lead to elimination of the largest proportion of paths, when they were deleted. The complete list paths are provided in Supplement 4.

**Figure 3:**
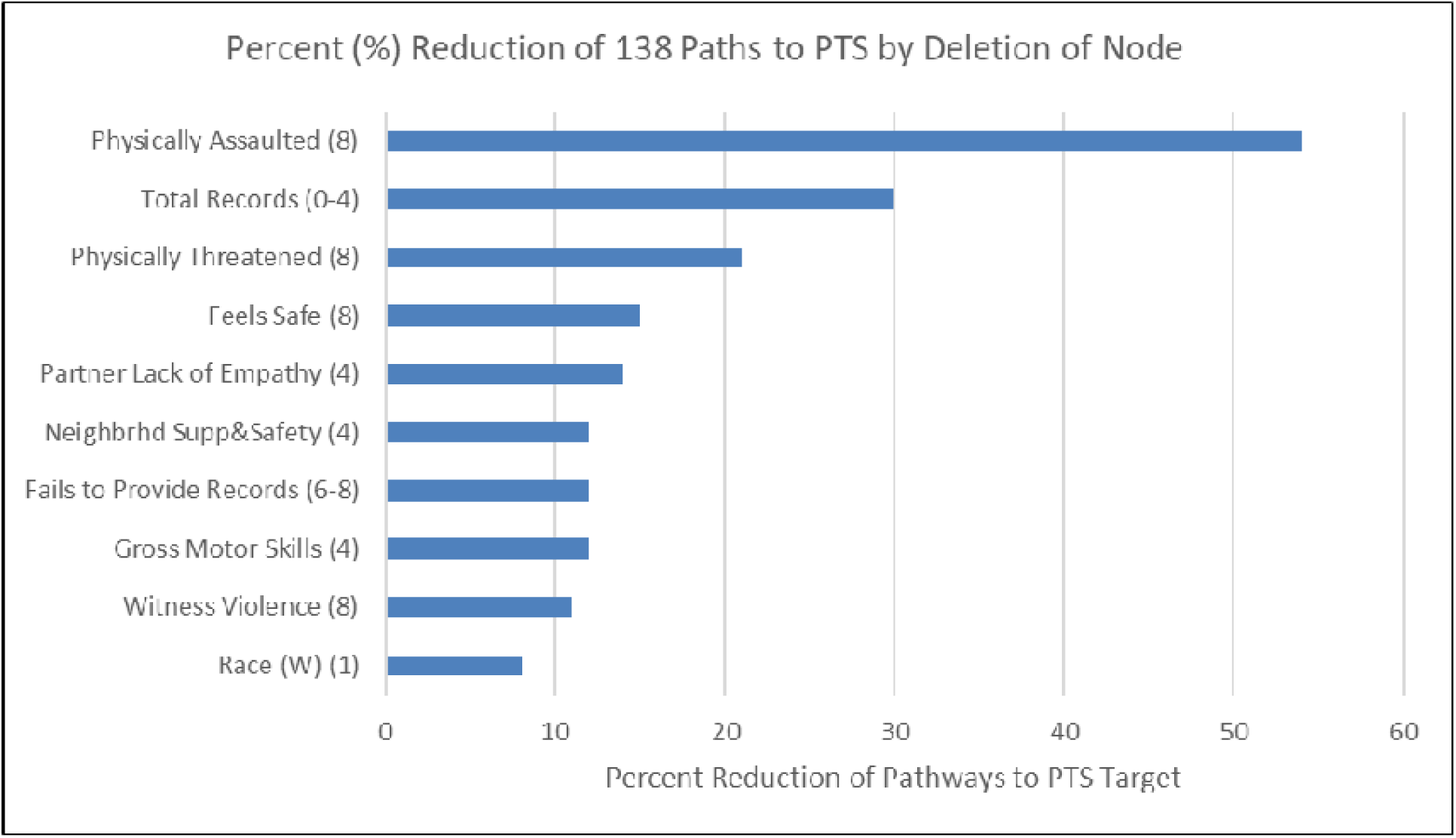
Rank order of variables with influence on greatest proportion of paths to PTS (time époque in parenthesis)

## Discussion

A Causal Network Model for PTS was revealed that included 4 direct causes, 2 direct effects, and 138 causal pathways. This model was determined with methods rarely applied in the field and so caution in interpretation of these results is warranted. In this context, the promise of CCD methods would be supported if the results are consistent-enough with previous literature, but still reveal novel findings.

Our results show strong consistency with previous literature with findings that different forms of trauma (e.g. witness violence, physical assault) combined with the child’s sense of safety, are related to PTS. Previous work could not infer causal relations between trauma exposure variables and PTS. CCD methods determined that such relations are unconfounded by a broad range of possible sources of influence. Moreover, the resulting causal network shown in Figure 2, indicates how these trauma exposure/safety experience variables may inform understandings of PTS and reveal opportunities for intervention. For example, feeling of safety at age 8 is related to expression of socially desireable responses, and such expression is also influenced by level of peer acceptance earlier in development (age 4-6), and to physical assault at age 8. Social desireabilty at age 8, in turn, influences one of the primary downstream effects of PTS – negative self image – combined with feeling unsafe, level of adaptive function, and exposure to domestic violence earlier in life (age 4-6). Accordingly, this causal model identifies pathways to a critical developmental problem (negative self image) involving the complex relations between trauma exposure, feelings of safety, PTS, adaptive function, peer acceptance and socially desireable responses. These findings highlight the need to understand a child’s experience of belonging/acceptance within their social groups to prevent a significant developmental consequence of PTS.

Another observed effect of PTS was exposure to severe assault by a non-caregiver at age 8. Such a finding may seem surprising: Why would PTS lead to severe assault by non-caregivers? This finding may, however, be understood through the natural role of parents as protectors of their children. A child with distress from PTS would naturally try to elicit help and protection from their parents/caregivers: but this observed pathway is also related to severe emotional maltreatment and maltreatment related to substance abuse. This finding may highlight the child with PTS’s vulnerability to elicit help from adults who do not have a caregiving role, because their parents/caregivers are unavailable to provide such help. Not only would such parents/caregivers be unavailable/unable to provide help for distress, they would also be unavailable/unable to provide protection from non-caregiving adults who their children approach for help, should protection be needed. Future research should try to clarify and replicate this finding, particularly because bootstrapping analysis revealed this relationship may be influenced by sampling variation.

The variables included in Figure 3 provide complementary information to those shown in Figure 2. Results indicate that PTS is multiply determined, but some of the identified causes are not directly remediable after they occur (e.g. witness violence, physical assault). Our approach to modeling intervention on all observed causal variables enables the identification of those with broadest impact on causal paths that produce PTS. A diverse set of variables were identified that, in general, describe processes earlier in life than the direct causal variables, supporting the notion that these variables would represent more pervasive influences on PTS. Such broadly influential variables (and means of their remediation) include feelings of safety at age 8 (help child feel safer), partner of primary caregiver’s lack of empathy at age 4 (couples therapy, individual therapy for primary caregiver), and child’s motor skills at age 4 (early developmental intervention). Should such findings be replicated, they indicate areas to assess for possibile preventative intervention.

There are specific reasons for caution related to some of our results. As shown in Figure 2, edges between several variables were observed in less than 30% of bootstrapped samples. Although the finding of a causal edge in any of the bootstrapped samples is valid, the appearance of an edge in a small number of such samples points to sensitivity of the edge to sampling variation, indicating greater need to replicate such a finding. Additionally, PCCDP cannot (at its present configuration), rule out the possibility that an unmeasured hidden common cause may explain an observed causal relationship. At present, the methods employed enable consideration of far more variables (252) than can possibly be considered using conventional methods, and common causes from these 252 measured variables can safely be excluded. Our analyses do not, however, rule out the possibility of confounding from unmeasured variables. It is in principle possible to detect such effects by using additional methodologies,^21,49^ and we intend to explore extensions to PCCDP for this purpose.

As stated, we believe caution is warranted in accepting our results. Correspondingly, we also believe caution is warranted in simply dismissing our results, because they are based on methods that deviate from the exclusive paradigm for causal inference (i.e. experimental research) used by the field for decades. We applied these methods in the present study to address a central limitation of this paradigm which - paradoxically - has served to limit the quality the causal knowledge-based held in the field, because: 1. In most cases, human experiments on etiology cannot be conducted and, thus, most published studies pertaining to etiology use correlational methods: yet the findings from such studies are typically discussed in causal terms, and 2. The main risk for false discovery in research using correlational methods concerns hidden common causes (from both unmeasured variables and measured variables unexamined as possible common causes) that serve to confound the observed relationship between a presumed causal variable and a presumed effect. If only a small proportion of the actual causal factors for a given effect (i.e. an outcome) are considered within a given study (likely for most correlational studies in psychiatry), the likelihood of confounding from hidden common causes is very high for that study.

It is imperative to advance knowledge on causal factors for PTS in maltreated children, so that effective interventions for these children may be developed and implemented. The present study is submitted in order to contribute to such much needed advances.

## Data Availability

All data produced in the present study are available upon reasonable request to the authors.

