## Supplement 1 for "Computational Causal Discovery for Posttraumatic Stress and Negative Self Image in Young Maltreated Children"

A Non-technical Overview of Computational Causal Discovery

The adoption of causal discovery methods that lie outside the randomized experimental framework require a change of paradigm. Within the current, conventional research paradigm in the field, knowledge on causation can only come from experiments, and therefore claims about causal inference from observational data will typically arouse a great deal of skepticism. It is beyond the scope of this article to review in detail the literature on causal inference and to compare and contrast how this inference can come from experimental research and from non-experimental, observational research. These matters have been extensively reviewed elsewhere, including both rigorous mathematical proofs justifying these claims and extensive empirical validation studies using these methods to discover causes using gold standard causal discovery data sets where true causes are known (refs). The related technical literature as well as applications studies date as early as the 1950s and have produced theoretically robust and empirically successful results, that have had major implication in diverse fields such as economics and econometrics, computational biology, genomic medicine, cancer genomics, and others. For the benefit of the mental disorders researchers who may not be familiar with such methodologies, we provide the following brief non-technical overview on how causation may be inferred from observational data.

A. **Operational definition of causality.** We start be giving a definition that explicitly or implicitly is used by all state of the art causal discovery algorithms and is consistent with what is considered to be causal in biomedicine:

Definition 1 (Operational criterion for causation): Assume that a variable A can be manipulated by a hypothetical experimenter to take values denoted as a_i_. Assume also that the experimenter can manipulate A (e.g., give a drug to a patient or not, where A sstabnds for type of treatment received). We denote the manipulation of A by the experimenter to take value aj, as: do(A=aj). If the experimenter assigns values to A according to a uniformly random distribution over values of A, and then observes P(B|do(A = a_i_))≠P(B|do(A = a_j_)) for some i and j, (and within a time window *dt*), then variable A is a cause of variable B (within *dt*). (Pearl, 2000).

Definition 2 (Direct and indirect causation): Assume that a variable A is a cause of variable B according to the operational criterion for causation in definition 1. A is an indirect cause for B with respect to a set of variables V, if and only if A is not a cause of B for some assignment (by manipulation) of values of V - {A, B}, otherwise A is a direct cause of B*.* Variables representing direct causes serve to *mediate* the relationship between an indirect cause and the effect.

**B. Mapping of data to the causal process generating the data.** The existence of a set of causal relationships of the type “A causes B” in the vast majority of distributions (so-called “Faithful” distributions) guarantee that specific conditional dependencies and independencies will be observed in the data in a way that directly corresponds to the causal mechanisms of the process that generates the data. For example, consider the situation that A directly causes B, B directly causes C and no other direct causal relationship exists among them. In this case all variables are mutually correlated (or broadly speaking dependent, if correlation is not the most appropriate form of measured association). Also, A will become independent of C if we condition on B (i.e., knowing the values of B blocks the association/dependency/information transfer between A and C). This is an application of the **Causal Markov Condition (CMC)**, a distributional assumption that allows us to infer all statistical independecies observed in data if we know the causal process generating the data. The distributional assumption of **Faithfulness Condition (FC)** says that *all* indepedencies in the data are represented by the causal graph representing the causal process combined with the CMC. If the CMC and the FC hold, it follows that all dependencies and independencies in the data correspond precisely to (i.e., form a perfect map of) the true causal structure that generated the data. In other words an algorithm (or analyst if the number of variable sis really small) can measure dependencies and independencies in the data and correctly infer the existence or non-existenceof precise direct and indirect causality without conducting experiments. The Combination of CMC+FC constitutes the aforementioned. distribution assumption of Faithfulness. How common are faithful distributions? The theoretical answer is that the non-faithful distrbutions among all possible theoretical ones are exceedingly rare (Lebesque measure 0) (REF: Meek). In practice we know few cases where faithfulness is violated and for many of these cases we have special algorithms that can cope (e.g., multiplicity (REF: Statnikov).

**C. Completeness of discovery procedures.** Can all direct and indirect causal relationships be revealed by the causal discovery algorithms? The answer is no, there are cases that parts of the causal process cannot be deciphered without experimentation because more than one conflicting causal processes are consistent with the data. Consider for example, the case where we measure only 2 variables , and have no temporal information about their order. The causal algorithm will infer that A causes B or B causes A. However, the advantage that the causal discovery algorithms give us is that the number of experiments we need conduct is commonly a tiny fraction of the experiements that would be needed if we we would use exclusively experimentation. Newer generations of hybrid causal active learning algorithms (e.g., ODLP REF: Ma, Statnikov et al)) can actively minimize the number of experiments that a researcher need perform.

D. **Direction of causality.** Whenever partial or complete information exists about the temporal order of variables, the causal discovery algorithms can incorporate it and learn the correct structure easier. When temporal order is unknown, however, its discovery from data is still possible by utilizing a variety of approaches, such as detecting so-called “Y structures” and propagating directionalities in the causal network, or using mnethod epxloting the asymmetry of conditional dependence distributions in nature, etc. (REF: ***).

**E. Hidden variables.** When not all variables are observed, hidden variables may exist that act as unmeasured confounders. Special extensions of the methodology described above allow for the discovery of conditional dependency and indepednece “signatures” in observational data that correspond to specific hidden variable structures. Protoypical algorithhms in the category (e.g., FCI, IC* (REF ***)) will be able to resolve some of the hidden variables, whereas the remaining ones will require experimentation to be resolved. Everything else being equal, the more variables we have observed, the more likely that hidden variables will be resolved.

**F. Estimating quantitative effects.** Everything we covered so far addresses qualitative discovery of the type “A directly causes B”. The next step is quantitative discovery whereas we estimate that “1 unit change of A will result in X units of change in B (everything else being constant)”. This type of inference is given to use by the apparatus of “Do-Calculous” (REF Pearl). The do calculous specifies the variables that we need to condition on a regression (or other appropriate quantitative model) such that the estimated coeffient for the effect or A on B is the true causal effect. The Do-calculous conditioning variables must be taken from a correct causal graph of the process that generates the data, thefore the approach is: infer qualitative causal graph🡪 infer conditioning variables from the graph🡪estimate causal effects. Note that simply putting a number of variables we believe or suspect are confounders of A and B will NOT in genral give as a correct estimate of the true causal effect.

G**. Modeling the effects of hidden variables.** Under conditions it is even possible to quantitatively estimate the effects of unmeasured variables on measured ones. Se for example REF Kummerfeld et al.

H. **The need for advanced computation.** Because when the number of causal models and depednencies and independencies as a function of the number of variables is worst case exponential, all the analyses required by causal discvery methods require not just the use of computers but the development or smart algorithms that reduce the number of required computations by employing heuristics, and other scalability methods. One such approach is local methods and local-to-global methods (REF: aliferis et al jmlr 2010a, 2010b).

**I. Relationship with predictive modeling.** Newer predicitve modeling algorithms have had tremendous success acorss many fields in recent years. Often predictive models are abused by being misinterpreted causally. This is a big pitfall that needs be avoided (REF Aliferis 2010a). There is however a natural connection between predictive modeling, feature selection and local causal discovery in the family of Markov Boundary algortihms. These algorithms combine local causal properties with optimal predictivity and dimensionality reduction (REF: Aliferis 2010a,b). In our work we make extensive use of such methods.

J**. What about prior knowledge?**. A not uncommon set of related criticisms of computational discovery is that of being “hypothesis free”, ignoring prior literature, not leveraging domain theory , being inconsistent with domain theory, and being a “fishing expedition”. These criticisms may or may not be valid depending on how investigators choose to use the methods, but they are not inherent weaknesses. If investigators collect data, run algorithms on data, and interpret models without any consideration of prior work in the field then the criticisms are very valid. If, however, the definition of the problem to be examined, the sampling and encoding of data, the configuration of algorithms to take into account robust prior knowledge, and the use of prior knowledge to resolve equivalent models with conflicting results is conducted on a foundation of solid prior knowledge, then the application of the methods will yield better results and the criticisms will not be applicable.

**K. Acyclical versus cyclical processes.** The vast majority of methods for causal discovery form observationa data utilizes **Directed Acyclic Graphs (DAGs)** and presupposes acyclic processes. These assumptions may be violated in certain fields and this generally addressed by modified methods that infer causal relationships in systems with feedback loops in quilibria, or in systems with feedback loops that are untangled over time by using temporal data (REF***). Even if a system has cyclical relationships, and analysis method assume acyclicity, this will typically manifest as unresolved flagged relationships that require further investigation by the researcher.
